# Month-to-month all-cause mortality forecasting: A method to rapidly detect changes in seasonal patterns

**DOI:** 10.1101/2023.02.07.23285581

**Authors:** Ainhoa-Elena Léger, Rizzi Silvia

## Abstract

**Background:** Short-term forecasts of all-cause mortality are used retrospectively to estimate the baseline mortality and to obtain excess death after mortality shocks, such as heatwaves and pandemics, have occurred. In this study we propose a flexible method to forecast all-cause mortality in real-time and to rapidly identify short-term changes in all-cause mortality seasonal patterns within an epidemiological year.

**Methods:** We use all-cause monthly death counts and ratios of death counts between adjacent months as inputs. The ratio between one month (earlier month) and the consecutive month (later month) is called later/earlier ratio. We forecast the deaths one-month-ahead based on their proportion to the previous month, defined by the average later/earlier ratio over the preceding years. We provide forecasting intervals by way of a bootstrapping procedure.

**Results:** The method is applied to monthly mortality data for Denmark, France, Spain, and Sweden from 2012 through 2022. Over the epidemiological years before COVID-19, the method captures the variations in winter and summer mortality peaks. The results reflect the synchrony of COVID-19 waves and the corresponding mortality burdens in the four analyzed countries. The forecasts show a higher level of accuracy compared to traditional models for short-term forecasting, i.e., 5-year-average method and Serfling model.

**Conclusion:** The method proposed is attractive for health researchers and governmental offices to aid public health responses, because it uses minimal input data, i.e., monthly all-cause mortality data, which are timely available and comparable across countries.

**Keymessages:**

- What is already known on this topic: There is a lack of methods to forecast all-cause mortality in the short-term in a timely or near real-time manner.
- What this study adds: The method that we propose forecasts all-cause mortality one month ahead assuming a seasonal mortality structure and adjusting it to the level of mortality of the epidemic year. These aspects make the method suitable for forecasting in a timely manner also during mortality shocks, such as the current COVID-19 pandemic.
- How this study might affect research, practice or policy: The forecasts obtained with the proposed method detects changes in all-cause mortality patterns in a timely manner and can be used to aid public health responses.

## Introduction

In temperate countries in the Northern Hemisphere, all-cause mortality exhibits a marked seasonality, with a winter peak driven by influenza-related mortality among the older population [1]. Variations occur from year to year in the magnitude and timing of the seasonal pattern as a result of the severity of the influenza type, waves of extreme temperature in the summer, or mortality shocks, such as pandemics caused by infectious diseases. These variations make the annual impact of mortality difficult to predict at the beginning of each season. Accurate and timely mortality forecasts are needed, to aid public health responses by informing key preparation and mitigation efforts.

Various types of short-term mortality forecasts, i.e., of some weeks or months, are established in the literature. All-cause mortality is usually used as the outcome variable because it is readily available in most countries, and the occurrence of influenza epidemics is known to be associated with excess mortality for all causes [2]. The simplest way to forecast mortality is the average number of deaths or the average mortality rates over preceding years – for instance five years. Modelling is commonly preferred because it controls for time-varying population size and age structures, and it directly extrapolates a secular trend and estimates seasonal variations. The traditional Serfling model [3] predicts the expected weekly deaths by means of a cyclical regression. Several extensions have been developed, e.g., modelling monthly mortality rates [4], or performing a Serfling-Poisson regression [5]. More recent approaches use Poisson regressions, e.g., the EuroMOMO model for monitoring excess mortality [6], or time-series methods, e.g., the ARIMA or seasonal ARIMA (SARIMA) model [2, 7].

These methods are designed to predict mortality due to seasonal epidemics in the absence of mortality shocks. In the case of a shock, these forecasts are used retrospectively, to assess whether mortality exceeded the expected level. The forecasts are interpreted as baseline mortality (i.e., counterfactual forecasts), and excess mortality is obtained by subtracting the forecasted baseline from the observed mortality. Excess mortality has long been analyzed to quantify the severity of an influenza season [8-11], the mortality burden of heatwaves [12-13] and previous pandemics, such as the 1918-19 Pandemic (H1N1 virus) [4]. Excess mortality became of major interest during the COVID-19 pandemic and was diffused by media outlets and in the scientific literature [14-16], to compare countries across time [17-20] and to evaluate the effect of policy interventions [21].

Another type of short-term mortality forecast is the realistic forecast, used to study the progression of infectious disease epidemics, for instance influenza outbreaks. The aim is to anticipate the evolution of the disease and be informative to the preparation and prevention of illnesses, hospitalizations, and deaths. Traditional mathematical models for these studies are compartmental models [22-24], e.g., the susceptible-infectious-recovered (SIR) and susceptible-exposed-infectious-recovered (SEIR) models. Other approaches use statistical methods, e.g., statistical regressions [25] and time-series models [26-27], or machine learning techniques [28]. These methods usually focus on several outcomes other than all-cause mortality, such as mortality and hospitalization from pneumonia, influenza, or respiratory diseases. In the context of the COVID-19 pandemic, they were used to model COVID-19 cases and deaths [29-33].

Short-term mortality forecasting focuses, therefore, either on all-cause mortality in absence of a shock (to compute excess mortality) or on cause-specific mortality related to the outbreak of infectious diseases. To our knowledge, at time of writing, no method exists to predict all-cause mortality in real time – both in regular epidemic years, where the bulk of mortality occurs in the winter – and during mortality shocks, when usual patterns are disrupted, and mortality increases unexpectedly. In this study, we combine the two aspects. The aim of the analysis is twofold: (1) to propose a simple and flexible method for forecasting all-cause monthly one month ahead; and (2) to investigate the validity of the forecasting method during both regular epidemic years and mortality shocks. The method estimates the deaths to be expected one month ahead, based on recent past seasonality and at current mortality levels. We show an application on historical forecasts in the last decade, including both non-COVID and COVID years, in Denmark, France, Spain, and Sweden.

## Methods

### Data

Since the beginning of the COVID-19 pandemic, national statistical offices started publishing timely all-cause weekly and monthly mortality data series (see Supplementary Materials, Supplementary data). We focused on Denmark, France, Spain, and Sweden because of the availability and the high quality of the data. Moreover, different population sizes allow for a robustness check of the method. We retrieved data on monthly deaths for the total population. We chose 2007 as the starting point because it is the first available year for Denmark.

The death counts were adjusted to be comparable across months of different lengths, and across leap years and non-leap years, following Nepomuceno et al. [34]. We assumed the average number of days in a month in both leap and non-leap years to be 30.44 days (365.25/12). The monthly death counts were multiplied by the ratio between 30.44 and the actual number of days in each month. To adjust for any difference in the annual total number of deaths after rescaling, we distributed the difference accordingly to the annual relative frequencies of the rescaled death counts. The relative frequencies were computed within the epidemic year, to account for the influenza season.

### The later/earlier method

An epidemic year (epi-year) is defined from July through June, covering part of two adjacent calendar years. The influenza season usually starts in October and ends in May, with a seasonal mortality peak between December and March. Winter seasonality and low summer mortality mean that deaths of adjacent months are highly positive correlated (see Supplementary Materials, Supplementary descriptive analyses) because an increase/decrease in one month is associated with an increase/decrease in the following month. For example, the correlation between the deaths in October and November ranges between 0.75 and 0.85 in the countries analyzed. January, February, and March exhibit a lower correlation because of the variability in the timing and the shape of the winter peak. For example, the correlation between deaths in January and February ranges between 0.55 and 0.64.

By extrapolating the relation between the current month (earlier month) and next month (later month), one can predict the death counts in the next month at any moment in the epi-year. This reasoning is based on the later/earlier method introduced by Rizzi and Vaupel to make counterfactual forecasts after a major shock, e.g., the first COVID-19 wave [35-36]. The real-time forecasting method that we propose uses two adjacent months, instead of two parts of the epi-year. The ratio of deaths between adjacent months is the later/earlier ratio.

Formally, if *D*_*i,j*_ is the number of deaths in the *i* − *th* month during the *j* − *th* epi-year, the later/earlier ratio between the *i* − *th* month and the (*i* + 1) − *th* month in the *j* − *th* epi-year, denoted by *υ*_*i,j*_, is given by

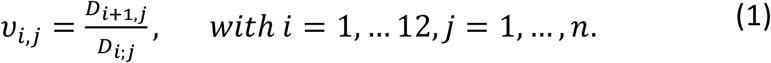

If the later/earlier ratio one month ahead is not known, one can assume that it equals the average later/earlier ratio of the previous years. The average later/earlier ratio 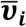 for the *i* − *th* month in the epi-years preceding the *j* − *th* epi-year is

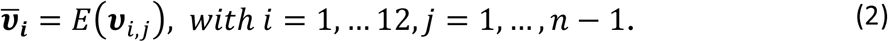

This assumption provides a short-term forecast of the expected deaths one month ahead. The values *υ*_*i,j*_, with *j =* 1, …, *n* − 1, can be checked for stationarity over previous years. If the series shows no trend, the deaths in the *i* + 1 − *th* month can be forecasted based on the deaths in the *i* − *th* month and on the average ratio 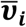 for the *i* − *th* month, according to the formula

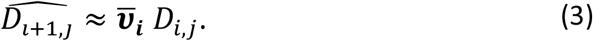

Let us suppose that we want to forecast the death counts one month ahead. Figure 1 shows, for illustrative purposes, the quantities used to forecast the death counts in epi-year 2013/14 (*j = n =* 6) in Spain. When there is an increase in the death counts, the corresponding later/earlier ratio is greater than 1; when there is a decrease, the later/earlier ratio is lower than 1. The average ratios of the previous five epi-years (*j =* 1, …, *n* − 1 *=* 1, …, 5) are greater than 1 from October through January, and lower than 1 in the remaining months. We multiply the death counts by the average later/earlier ratios to obtain the expected number of deaths in the next month.

**Fig. 1.**
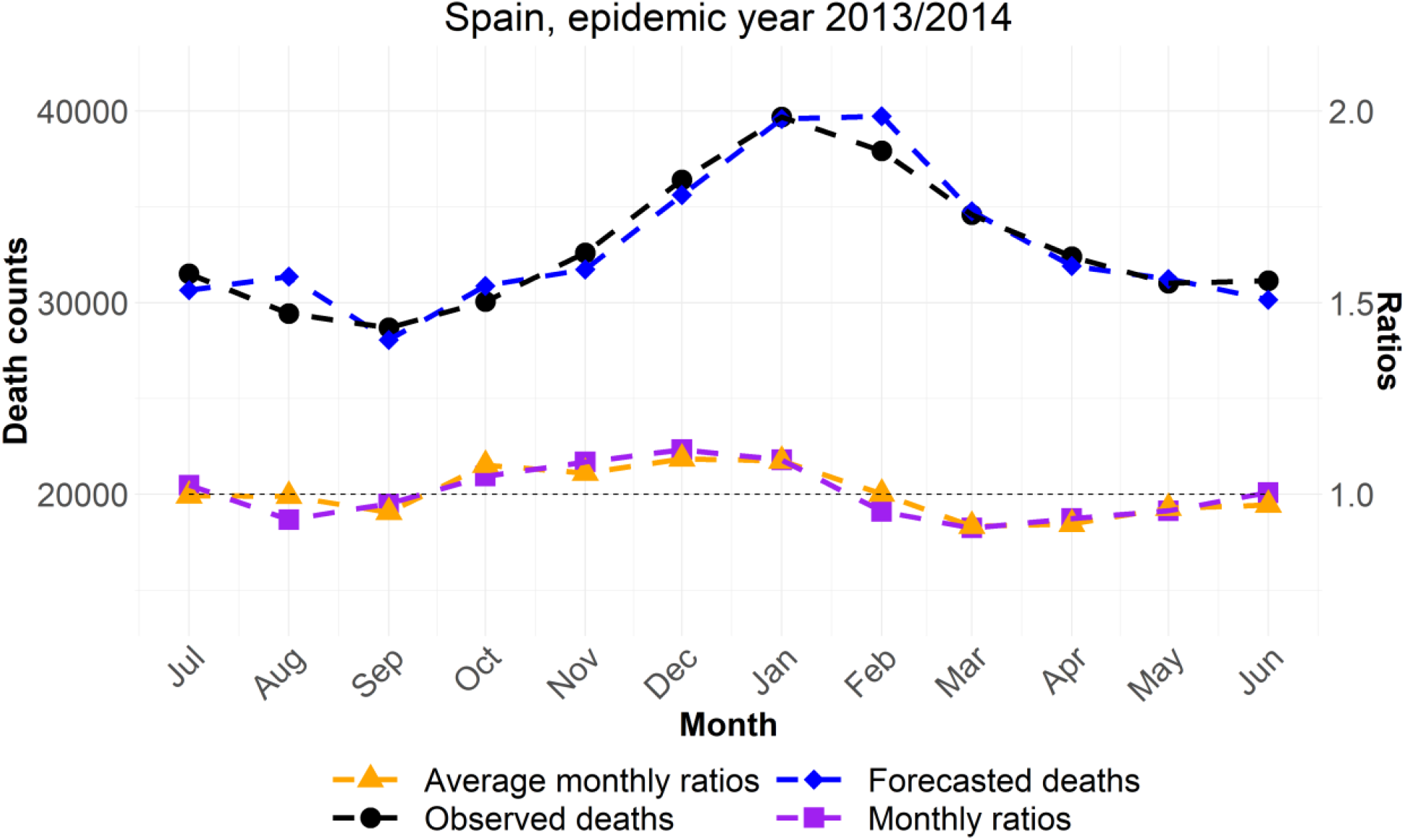
Illustration of the input data: Monthly death counts (black circles) and average later/earlier ratios (purple squares) compared to monthly ratios (orange triangles); and output: Forecast of the monthly death counts (blue diamonds) of the method. Source: Own elaboration.

### Prediction intervals

We computed the prediction intervals for the forecasts considering the two sources of variability. The first source comes from the uncertainty in the later/earlier ratios, i.e., from the assumption that the ratios equal the average ratios in the previous years. We drew 10,000 simulated ratios from the series of ratios through a bootstrapping procedure and used them in Equation 2 to compute 10,000 expected deaths one month ahead. The second source comes from the observed deaths. We drew 10,000 death counts from a Poisson distribution with the mean equal to expected deaths one month ahead. Empirical 95% prediction intervals were computed as the 2.5th and 97.5th percentiles of the Poisson distribution.

## Results

### Series of monthly death ratios

The later/earlier method assumes that the later/earlier ratios do not show any trend in the short term. The series of later/earlier ratios for Denmark, France, Spain, and Sweden revealed a considerable regularity, i.e., constant means and small standard deviations (see Supplementary Materials, Supplementary results). The constant averages and small coefficients of variation – lower than 9% – indicate a discrete correlation between the deaths in two adjacent months. The series are stationary via the Ljung-Box and the Kwiatkowski-Phillips-Schmidt-Shin tests [37]. A sensitivity analysis excluding COVID-19 epi-years (see Supplementary Materials, Supplementary sensitivity analysis) showed little effect on the average later/earlier ratios.

### Monthly forecasts with the later/earlier method

The later/earlier method produces realistic one-month-ahead forecasts. We applied it to forecast monthly mortality from July 2012 through December 2021 (July 2022 for France). The average later/earlier ratios were computed on the previous five epi-years, starting from 2007/08. Figure 2 illustrates the results for the four countries analyzed. The method is reactive to changes in mortality within and across epi-years because: a) it assumes a seasonal mortality structure in the epi-year given by the average later/earlier ratios (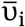 with i *=* 1, … 12 in Equation 2) and b) at the same time it adjusts the level of mortality within an epi-year using the actual observed deaths of the previous month (D_i,j_ with i *=* 1, … 12, j *=* 1, …, 5 in Equation 2).

**Fig. 2.**
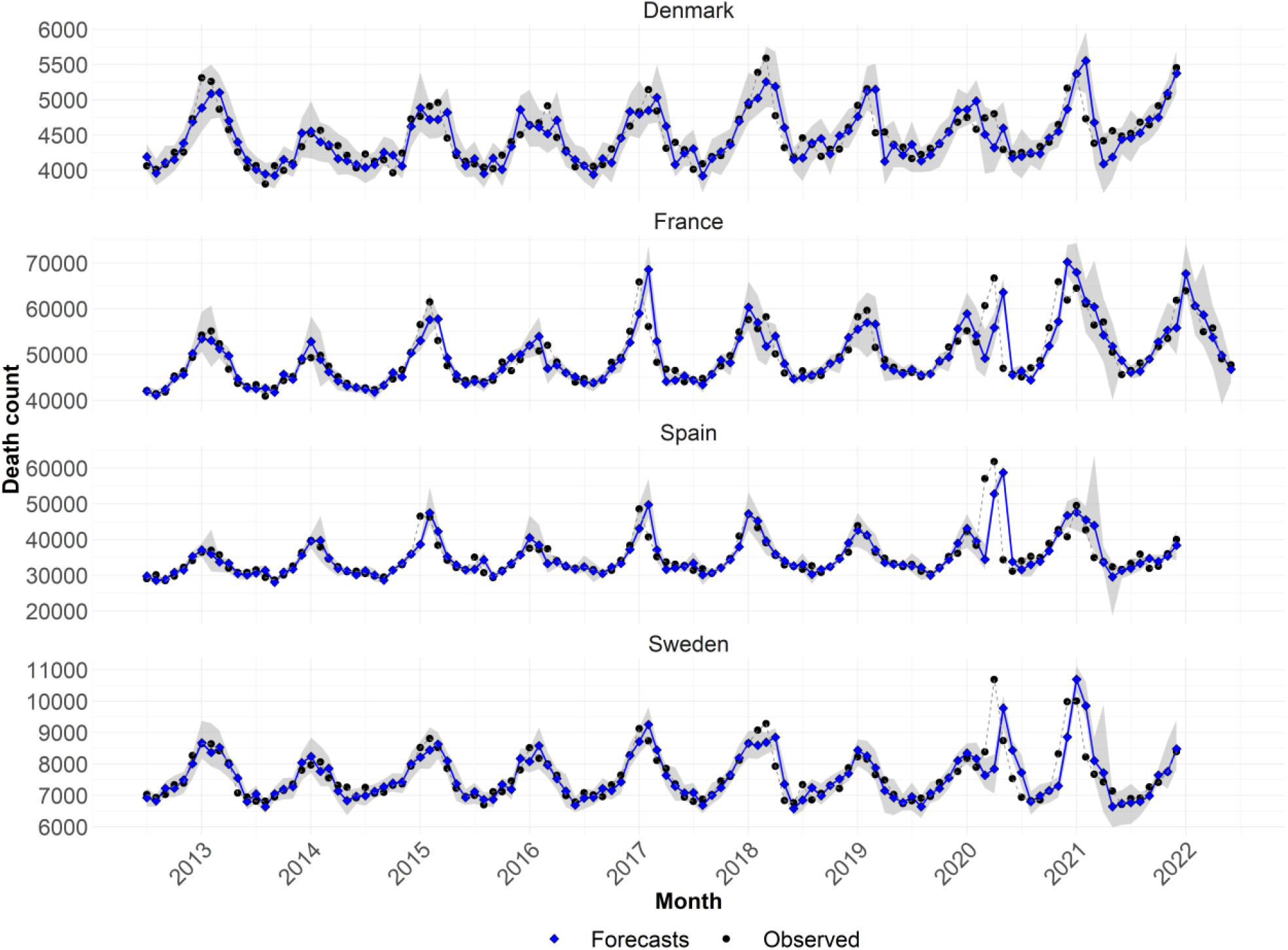
One-month-ahead forecasts (solid blue line with overlaid blue diamonds) with the later/earlier method starting from epi-year 2012/13 through epi-year 2021/22 in Denmark, France, Spain, and Sweden vs. observed death counts (dashed black line with overlaid black dots), and 95% prediction intervals via bootstrapping procedure (gray shaded area). Source: Own elaboration.

For example, the method captures the higher level of winter mortality in 2012/13 and the lower winter level in 2013/14 in Denmark, due to different types of viruses and transmission modes. The season 2012/13 witnessed a long period of high influenza activity dominated by A(H3N2), while influenza activity and mortality were low in the A(H1N1) dominated 2013/14 season [11]. A season with predominant influenza A(H3N2) has higher mortality impact than a season with predominant influenza A(H1N1) or a season with low influenza A transmission. As a second example, the method forecasts the higher seasonality peaks in the winters 2014/15 and 2016/17 in France and Spain. These seasons were characterized by a high influenza activity of A(H3N2) viruses, the circulation of variants of the virus and a reduction in the effectiveness of the flu vaccine [9].

Our forecasts reflect the synchrony of the three main COVID-19 waves through 2020 and 2021 [38] and the different mortality burdens in the four analyzed countries. A consistent excess mortality –large in Sweden and Spain, medium in France [17-20, 39-41]– was reported for the first wave (mid-February 2020 through end of May 2020). In the second wave (autumn 2020 through March 2021) and third wave (from the latter half of 2021 and on-going by the end of 2021), France experienced a similar toll to that of the first wave, whereas Spain and Sweden experienced lower tolls but lasting for many weeks [39].

The monthly prediction intervals are not equally affected by the overall seasonality. They are wider in correspondence of the winter peak, where variability in the observed deaths is higher. They are also wider for Denmark and Sweden, which have smaller populations and fewer deaths. Prediction intervals reflect mortality changes in preceding years. For instance, the uncertainty in the estimates for the months from March to June 2021 in France is greater because of the COVID-19 wave in 2020.

### Comparison with alternative methods

To assess the accuracy of the forecasts, the later/earlier method was compared with the 5-year-average method and the quasi-Poisson Serfling model (see Supplementary Materials, Supplementary methods). We chose these well-established methods for short-term mortality forecasting because they allow the same input data as the later/earlier method, i.e., a rolling window of 5 epi-years of all-cause mortality data. Figure 3 illustrates the respective monthly forecasts when each of the three methods are applied.

**Fig. 3.**
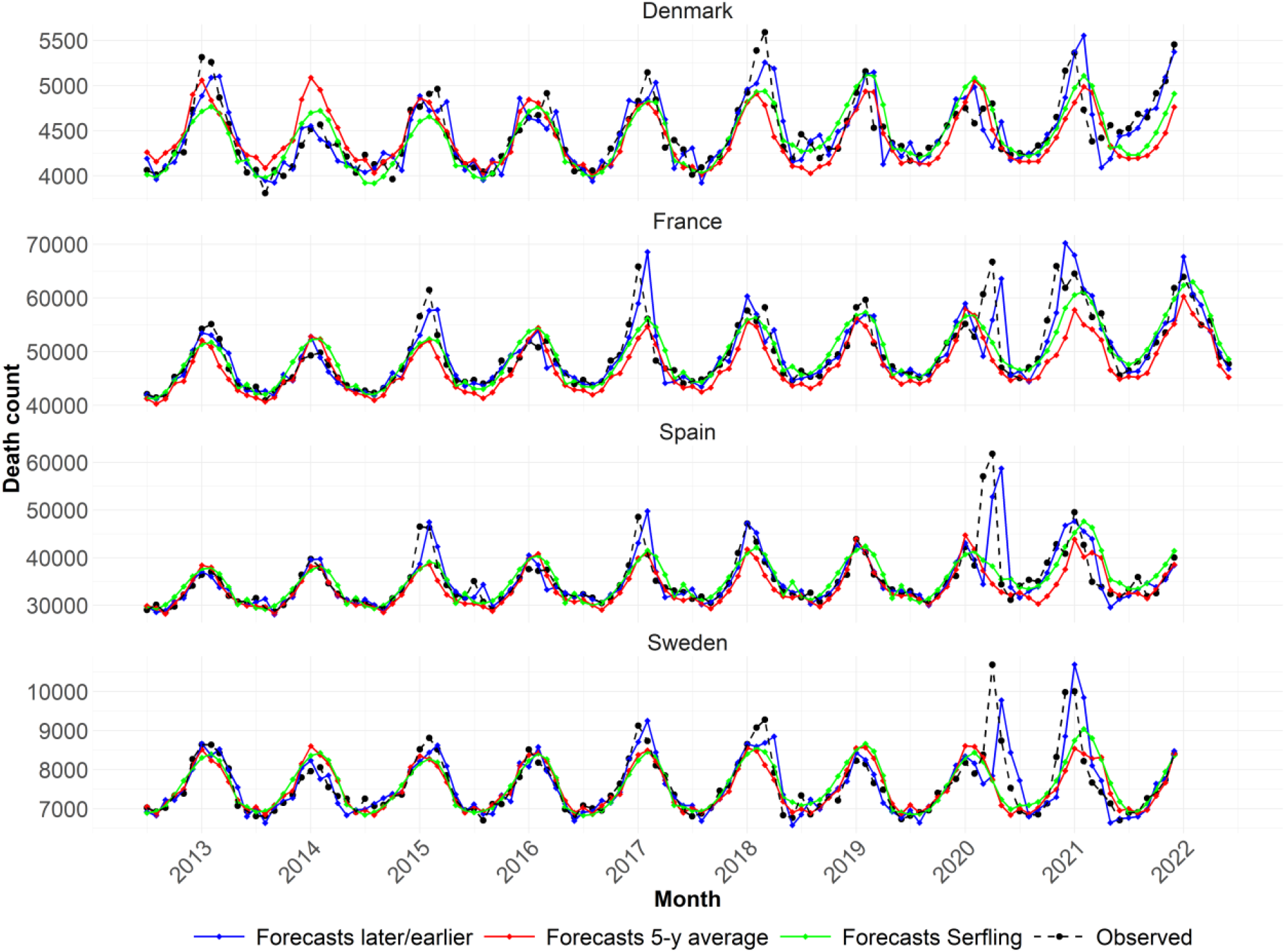
Observed death counts (black dots and dashed line); Forecasts with the later/earlier method (blue diamonds and solid line), 5-year-average method (red diamonds and solid line), and Serfling model (green diamonds and solid line) starting from epi-year 2012/13 through epi-year 2021/22 in Denmark, France, Spain, and Sweden. Source: Own elaboration.

The later/earlier method captures better the seasonality than do the other two methods. For example, the lower level of mortality in Denmark in the winter of 2014 is forecasted by the later/earlier method, while the 5-year-average and the quasi-Poisson Serfling predict a considerably higher level. In France and Spain in 2014/15 and 2016/17, the seasonality is also not captured by the 5-year-average and the quasi-

Poisson Serfling. The later/earlier method turned out to be the only method that forecasted the mortality shock in March and April 2020, albeit with one month of delay due to the sudden sharp mortality increase. The delay could be avoided by using a time window finer than a month. After the first COVID-19 wave, the later/earlier method provides better predictions than the 5-year-average and the quasi-Poisson Serfling because it is more flexible in relation to the higher winter mortality in 2020/21 in the four countries, and in Denmark in 2021/22.

To evaluate the goodness of the forecasts, we computed the root mean squared error (RMSE) and the mean absolute percentage error (MAPE) of the forecasts of the three methods. The later/earlier method proved to be more accurate and less biased, followed by the 5-year-average and the quasi-Poisson Serfling. The RMSE, displayed in Table 1, is lower for the later/earlier method in 60.42% of the cases, followed by the 5-year-average (31.25%) and the quasi-Poisson Serfling (8.33%). The MAPE (Table 2) is lower for the later/earlier methods in 47.92% of the cases, followed by the 5-year-average (27.08%) and the quasi-Poisson Serfling (25%).

**Table 1:**
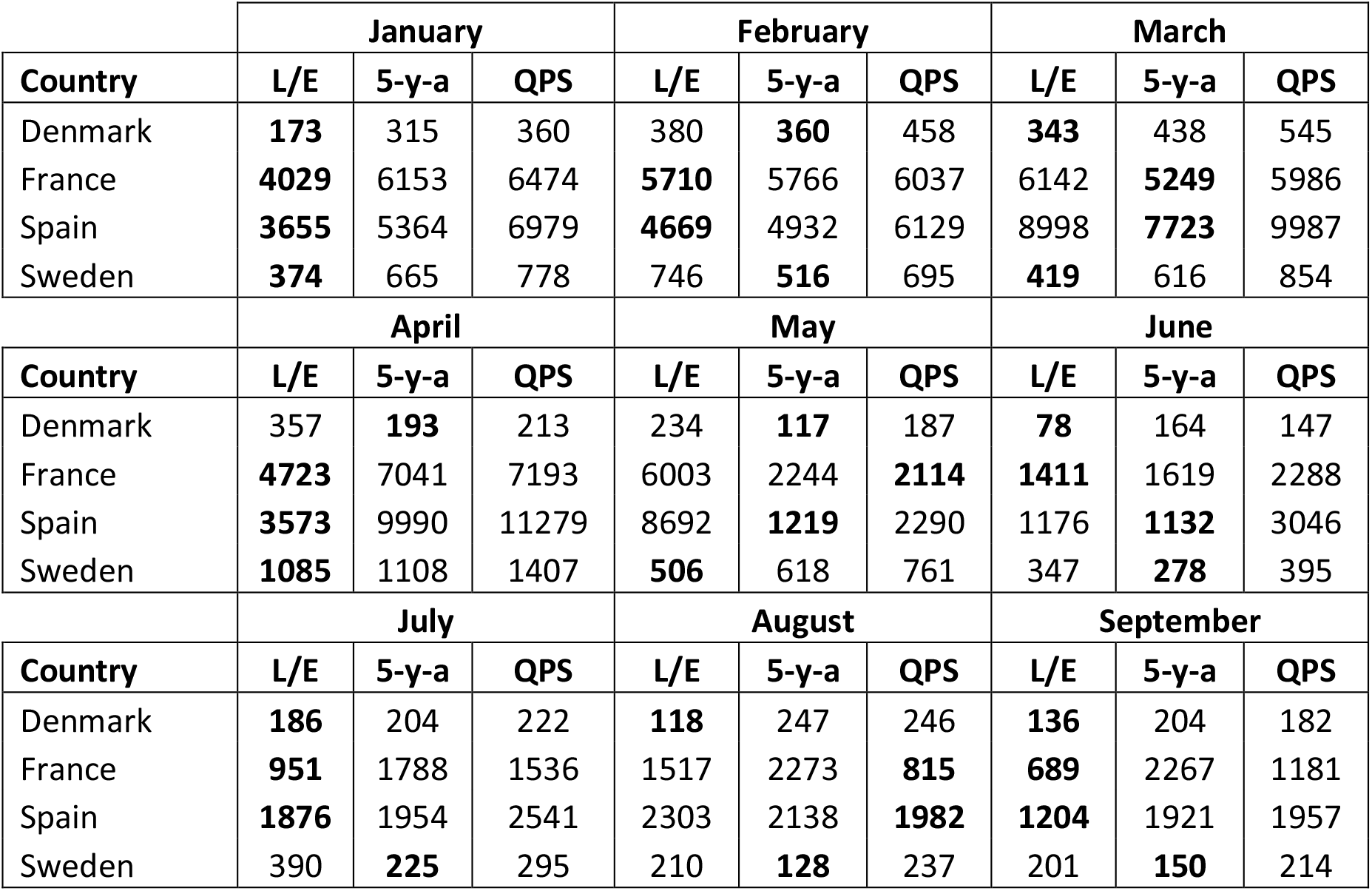
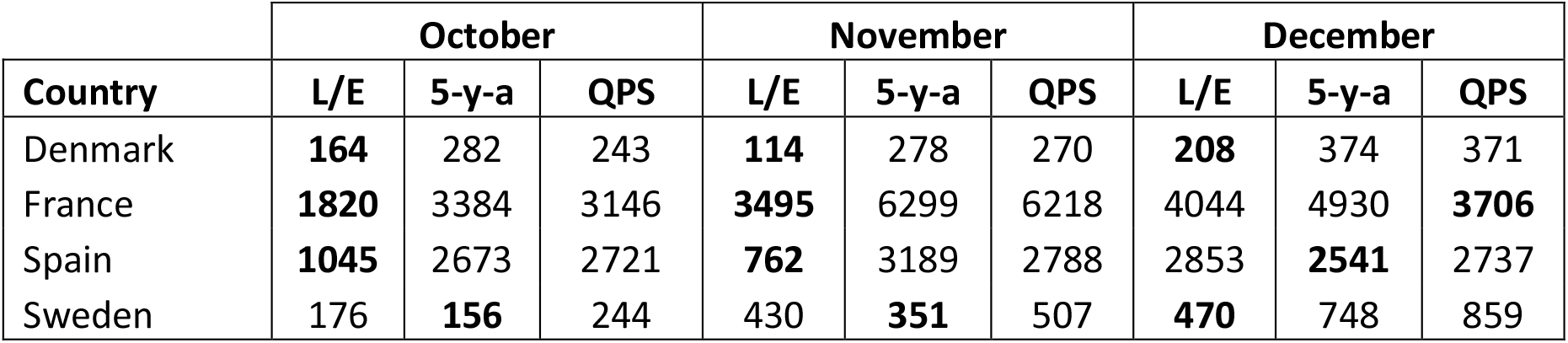
Comparison of the accuracy measure root mean square errors (RMSE) with the later/earlier ratio method (L/E), 5-year-average method (5-y-a), and the quasi-Poisson Serfling model (QPS) of the forecasts from epi-year 2012/13 through epi-year 2020/2021, by country. Bold numbers indicate the lowest and therefore best RMSE value between the three methods compared by country and month.

|  | January |  |  | February |  |  | March |  |  |
| --- | --- | --- | --- | --- | --- | --- | --- | --- | --- |
| Country | L/E | 5-y-a | QPS | L/E | 5-y-a | QPS | L/E | 5-y-a | QPS |
| Denmark | **173** | 315 | 360 | 380 | **360** | 458 | **343** | 438 | 545 |
| France | **4029** | 6153 | 6474 | **5710** | 5766 | 6037 | 6142 | **5249** | 5986 |
| Spain | **3655** | 5364 | 6979 | **4669** | 4932 | 6129 | 8998 | **7723** | 9987 |
| Sweden | **374** | 665 | 778 | 746 | **516** | 695 | **419** | 616 | 854 |
|  | April |  |  | May |  |  | June |  |  |
| Country | L/E | 5-y-a | QPS | L/E | 5-y-a | QPS | L/E | 5-y-a | QPS |
| Denmark | 357 | **193** | 213 | 234 | **117** | 187 | **78** | 164 | 147 |
| France | **4723** | 7041 | 7193 | 6003 | 2244 | **2114** | **1411** | 1619 | 2288 |
| Spain | **3573** | 9990 | 11279 | 8692 | **1219** | 2290 | 1176 | **1132** | 3046 |
| Sweden | **1085** | 1108 | 1407 | **506** | 618 | 761 | 347 | **278** | 395 |
|  | July |  |  | August |  |  | September |  |  |
| Country | L/E | 5-y-a | QPS | L/E | 5-y-a | QPS | L/E | 5-y-a | QPS |
| Denmark | **186** | 204 | 222 | **118** | 247 | 246 | **136** | 204 | 182 |
| France | **951** | 1788 | 1536 | 1517 | 2273 | **815** | **689** | 2267 | 1181 |
| Spain | **1876** | 1954 | 2541 | 2303 | 2138 | **1982** | **1204** | 1921 | 1957 |
| Sweden | 390 | **225** | 295 | 210 | **128** | 237 | 201 | **150** | 214 |

Table 1: Comparison of the accuracy measure root mean square errors (RMSE) with the later/earlier ratio method (L/E), 5-year-average method (5-y-a), and the quasi-Poisson Serfling model (QPS) of the forecasts from epi-year 2012/13 through epi-year 2020/2021, by country.
|  | October |  |  | November |  |  | December |  |  |
| --- | --- | --- | --- | --- | --- | --- | --- | --- | --- |
| Country | L/E | 5-y-a | QPS | L/E | 5-y-a | QPS | L/E | 5-y-a | QPS |
| Denmark | **164** | 282 | 243 | **114** | 278 | 270 | **208** | 374 | 371 |
| France | **1820** | 3384 | 3146 | **3495** | 6299 | 6218 | 4044 | 4930 | **3706** |
| Spain | **1045** | 2673 | 2721 | **762** | 3189 | 2788 | 2853 | **2541** | 2737 |
| Sweden | 176 | **156** | 244 | 430 | **351** | 507 | **470** | 748 | 859 |

**Table 2:** Comparison of the accuracy measure mean absolute percentage error (MAPE) with the later/earlier ratio method (L/E), 5-year-average method (5-y-a), and the quasi-Poisson Serfling model (QPS) of the forecasts from epi-year 2012/13 through epi-year 2020/2021, by country. Bold numbers indicate the lowest and therefore best MAPE value between the three methods compared by country and month.

|  | January |  |  | February |  |  | March |  |  |
| --- | --- | --- | --- | --- | --- | --- | --- | --- | --- |
| Country | L/E | 5-y-a | QPS | L/E | 5-y-a | QPS | L/E | 5-y-a | QPS |
| Denmark | **2.33** | 5.26 | 4.68 | **6.41** | 7.09 | 6.79 | **6.71** | 8.5 | 8.26 |
| France | **6.96** | 9.09 | 7.34 | 8.14 | 10.41 | **7.72** | 10.93 | **9.03** | 9.2 |
| Spain | **6.06** | 10.44 | 11.31 | **8.56** | 9.16 | 9.34 | 13.91 | **11.81** | 14.2 |
| Sweden | **3.87** | 6.18 | 5.99 | 7.69 | **6.33** | 6.81 | **4.61** | 6.96 | 7.95 |
|  | April |  |  | May |  |  | June |  |  |
| Country | L/E | 5-y-a | QPS | L/E | 5-y-a | QPS | L/E | 5-y-a | QPS |
| Denmark | 7.9 | 3.59 | **3.04** | 4.46 | **2.39** | 3.2 | **1.83** | 3.46 | 2.51 |
| France | 7.72 | 8.43 | **6.82** | 7.01 | 4.94 | **3.71** | **2.61** | 3.79 | 3.86 |
| Spain | **5.7** | 11.01 | 12.15 | 11.49 | **3.72** | 5.67 | **2.9** | 3.59 | 7.01 |
| Sweden | 7.68 | 7.56 | **7.36** | 5.7 | **4.98** | 5.54 | 3.22 | **2.79** | 3.84 |
|  | July |  |  | August |  |  | September |  |  |
| Country | L/E | 5-y-a | QPS | L/E | 5-y-a | QPS | L/E | 5-y-a | QPS |
| Denmark | 4.02 | 4.13 | **3.52** | **2.75** | 4.9 | 3.48 | 3.12 | 4.25 | **2.73** |
| France | **2.14** | 4.1 | 2.8 | 3.09 | 5.45 | **1.37** | **1.44** | 5.19 | 1.9 |
| Spain | 5.49 | **4.83** | 5.12 | 7.4 | 5.78 | **4.77** | **3.18** | 4.86 | 5.09 |
| Sweden | 5.27 | **3.03** | 3.23 | 2.88 | **1.53** | 2.54 | 2.9 | **1.97** | 2.18 |
|  | October |  |  | November |  |  | December |  |  |
| Country | L/E | 5-y-a | QPS | L/E | 5-y-a | QPS | L/E | 5-y-a | QPS |
| Denmark | 3.72 | 5.72 | **3.36** | **2.27** | 5.55 | 4.1 | **4.07** | 5.98 | 5.38 |
| France | **3.27** | 5.42 | 4.03 | **4.87** | 7.67 | 6.91 | 5.94 | 6.86 | **5.66** |
| Spain | **2.53** | 5.58 | 6.11 | **2.44** | 5.6 | 5.82 | 6.32 | 6.23 | **5.89** |
| Sweden | 2.33 | **2.07** | 2.81 | 4.15 | **3.77** | 5.24 | **4.4** | 5.27 | 5.07 |

## Discussion

The years 2020-22 will long be remembered as the years of the greatest pandemic to hit the world since the Spanish Flu of 1918-1920. For epidemiologists, public health researchers, and demographers, the pandemic has fostered many research questions. What methods and models are best suited to monitor and predict the severity of the current or future pandemics? And what lessons are to be learnt for non-pandemic mortality, or other kind of mortality shocks?

Mortality forecasting depends on data availability and data quality [29]. One limitation in infectious disease forecasting is that methods are very demanding in terms of information (cause mortality data, hospital data, mobility data, etc.). In theory-based models, e.g., compartmental models, estimates for case fatality rate, infection fatality rate, basic reproductive numbers (R0), and other key parameters essential in modelling, might be unavailable or inflated. Input data are usually available in highly specific settings, e.g., hospitals and care homes, leading to predictions produced on a small scale (3-32). Early during health shocks, registration systems are often not in place and limited data are available.

The few examples of all-cause mortality forecasts during severe mortality conditions are limited to seasonal influenza, such as the FluMOMO model, extending the EuroMOMO model to measure excess death [11]. However, these models require information other than mortality, usually data collection of indicators of temperature and influenza activity provided by surveillance systems, which are voluntary and insufficiently systematic and detailed [40]. Excess death is retrospective and does not permit a timely assessment. Furthermore, counterfactual estimates of baseline mortality might be outdated over several years.

Our study has proposed a novel method to produce realistic one-month-ahead forecasts of overall mortality, while requiring minimal input data and assumptions. The later/earlier method incorporates important features of mortality, learning from past seasonality of deaths and from the current yearly variable level of mortality. This ensures a flexible structure for forecasting in normal epidemic years, during and after a major shock, e.g., the COVID-19 pandemic. Moreover, the method can be extended to forecast other phenomena that vary periodically.

The main strength of the later/earlier method is that it requires minimal input. The choice of predicting all-cause mortality is particularly suitable for timely, realistic predictions. It avoids bias in causes of death registration (especially in the context of emerging pandemics), and issues of standardization of cause-of-death classification across countries and time. Furthermore, all-cause mortality data are usually available more readily, compared to, for instance, “COVID-19 death” to be confirmed by a testing procedure, or cause-specific deaths requiring a diagnosis.

The proposed forecasts rely on the assumption of stationarity of the series of later/earlier ratios. In our application, tests for stationarity proved that the assumption was met. If the assumption were not to hold, time series methods could be used to remove non-stationarity from the series of later/earlier ratios.

## Conclusion

Mortality monitoring is fundamental to health planning, risk assessment and public health action. Realistic and comparable forecasts are required to identify and react to changes in mortality patterns in a timely manner. In this study, we introduced and applied the later/earlier method to forecast monthly mortality. We applied it to four countries with different population sizes, and variation in pandemic phases and death tolls during the COVID-19 pandemic. Using this method, it proved possible to predict one month ahead the number of who would die. The forecasts produced showed to be competitive in terms of accuracy when compared to the well-established 5-year-average method and the quasi-Poisson Serfling regression.

The later/earlier method might serve statistical offices and surveillance systems in closely monitoring mortality progression, especially when little information is available, as it requires limited input data and no information on the spread of infectious diseases or cause of death. The method is flexible to changes in all-cause mortality because it assumes a seasonal mortality structure across epidemic years and forecasts adjusting it to the level of mortality within the given epidemic year. The later/earlier method might provide provisional estimates in the case of delays in registration. Given the pressing demand for vital statistics during the COVID-19 pandemic, statistical offices should continue to improve the release and the timing of reliable mortality data.

## Data Availability

Data from national statistical offices were used in this study. The full dataset and documentation can be downloaded from Statistics Denmark, INSEE, INE, and Statistics Sweden. The websites are provided in the Supplementary Materials.

https://www.statbank.dk/statbank5a/SelectVarVal/Define.asp?MainTable=DODDAG&PLanguage=1&PXSId=0&wsid=cftree

https://www.insee.fr/en/statistiques/serie/000436394

https://www.ine.es/dyngs/INEbase/es/operacion.htm?c=Estadistica_C&cid=1254736177008&menu=resultados&idp=1254735573002

https://www.statistikdatabasen.scb.se/pxweb/en/ssd/START__BE__BE0101__BE0101I/DodaManadReg/

## Acknowledgements

AL was supported by the AXA Research Fund “AXA Chair in Longevity Research” and SR by Rockwool Foundation “Excess Death Grant”.

James W. Vaupel provided us with insightful comments and suggestions, for which we are grateful.

## Statements & Declarations

### Author contributions

AL and SR designed the study. SR directed the implementation of the study. AL analyzed the data and graphically presented the results. AL and SR interpreted the findings, drafted the article, revised it critically, and approved the version to be published.

### Ethical approval

This research project did not require ethics approval as it used only macro data that are freely available online.

### Code availability

All statistical analysis were performed using R Software version 4.2.2. The R code for full replicability of the analysis of this paper will be made available in an online repository on GitHub.

