## Supplementary Materials for "Month-to-month all-cause mortality forecasting: A method to rapidly detect changes in seasonal patterns"

Journal of Epidemiology and Community Health

Supplementary Material

**Table of contents**

1. Supplementary Data

List of monthly data sources by country

2. Supplementary Descriptive Analyses

Correlation between death counts

Supplementary Figure S1. Correlation between deaths in consecutive months, year 2007 to 2022, France.

Supplementary Figure S2. Correlation between deaths in consecutive months, year 2007 to 2021, Spain.

Supplementary Figure S3. Correlation between deaths in consecutive months, year 2007 to 2021, Denmark.

Supplementary Figure S4. Correlation between deaths in consecutive months, year 2007 to 2021, Sweden.

3. Supplementary Methods

Competitive predictive methods

4. Supplementary Results

Series of later/earlier ratios

Supplementary Figure S5: Later/earlier ratios (black dots) and average later/earlier ratios (vertical black bars).

5. Supplementary Sensitivity Analysis

Sensitivity analysis of the series of later/earlier ratios

Supplementary Figure S6: Average later/earlier ratios with (vertical black bars) and without (vertical red bars) covid years, with a later period of one month and an earlier period of one month.

### 1. Supplementary Data

#### List of monthly data sources by country

**Denmark**. Statistics Denmark: Deaths by day of death and month of death (2007-2020). Available at https://www.statbank.dk/statbank5a/SelectVarVal/Define.asp?MainTable=DODDAG&PLanguage=1&PXSId=0&wsid=cftree (accessed on 03.11.2022)

Coverage: All death events registered in Denmark among its residents, regardless of their citizenship.

**France**. INSEE, L'Institut national de la statistique et des études économiques: Demography - Number of deaths - Metropolitan France, 1946-2021. Available at https://www.insee.fr/en/statistiques/serie/000436394 (accessed on 03.11.2022)

Coverage: The whole population of metropolitan France (excluding deaths in overseas territories).

**Spain**. Instituto Nacional de Estadística: Defunciones por edad, mes y sexo. Definitivos (1975-2020) and Provisionales (2021). Available at https://www.ine.es/dyngs/INEbase/es/operacion.htm?c=Estadistica_C&cid=1254736177008&menu=resultados&idp=1254735573002 (accessed on 03.11.2022)

Coverage: All the deaths that occurred in Spain.

**Sweden**. Statistics Sweden, SCB: Deaths per month by region, Region of birth, age and sex. 2000-2020. Available at https://www.statistikdatabasen.scb.se/pxweb/en/ssd/START__BE__BE0101__BE0101I/DodaManadReg/ (accessed on 03.11.2022)

Coverage: The whole resident population of Sweden, regardless of their citizenship.

#### 2. Supplementary Descriptive Analyses

##### Correlation between death counts

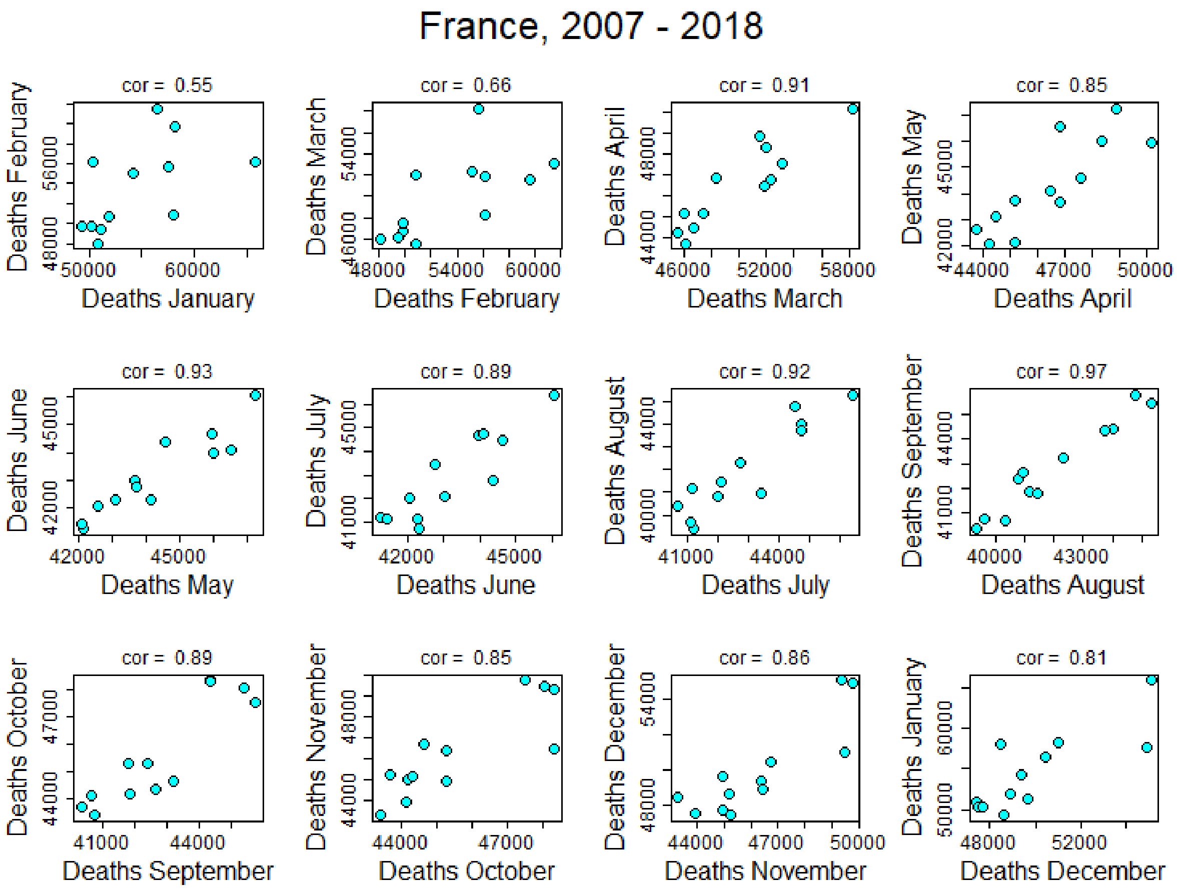


Supplementary Figure 1. Correlation between deaths in consecutive months, year 2007 to 2022, France. Source: Own elaboration.

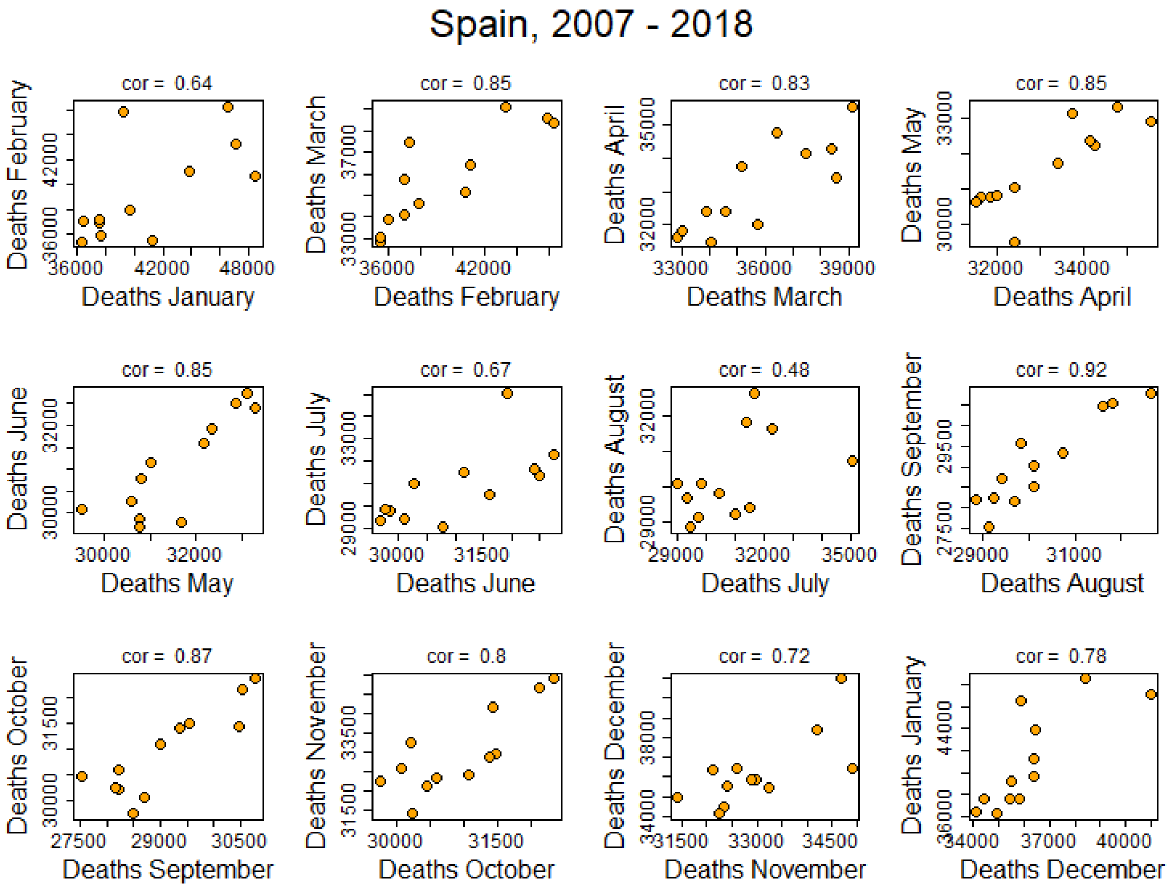


Supplementary Figure 2. Correlation between deaths in consecutive months, year 2007 to 2021, Spain. Source: Own elaboration.

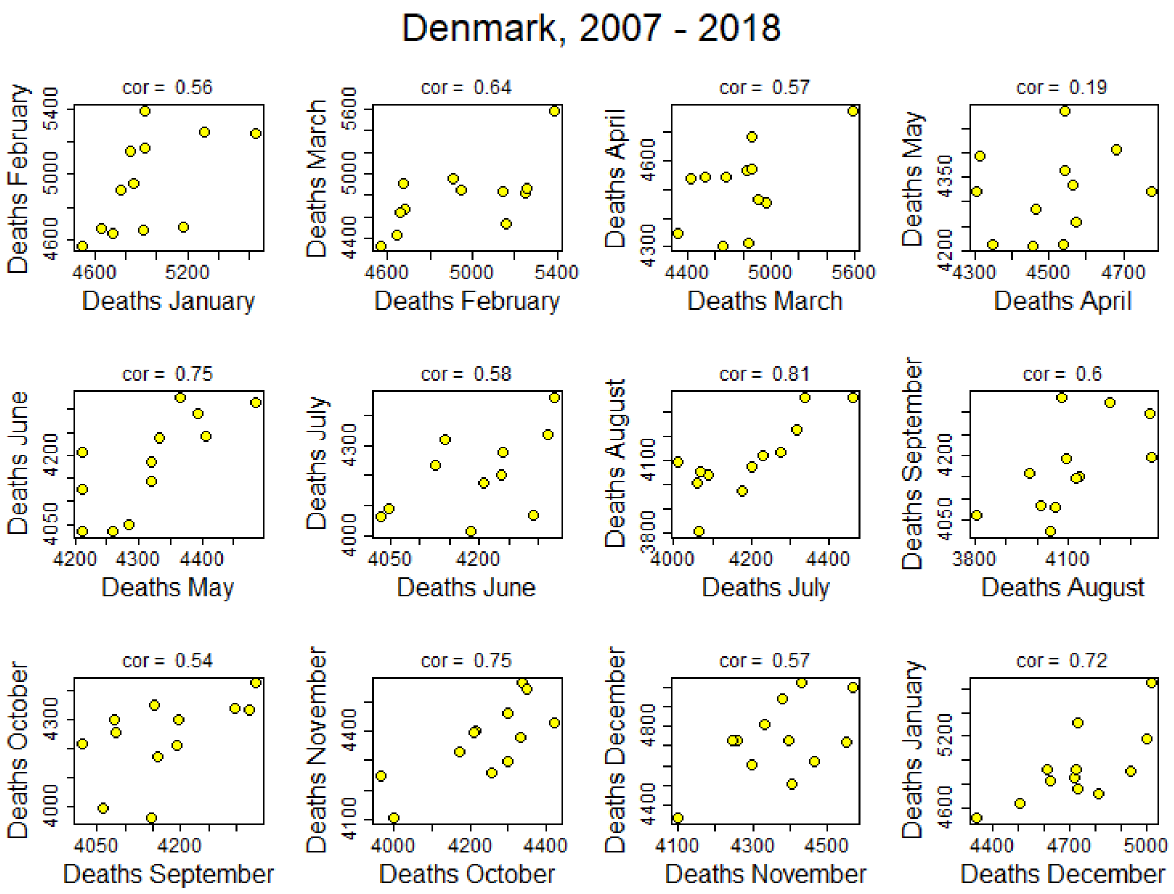


Supplementary Figure 3. Correlation between deaths in consecutive months, year 2007 to 2021, Denmark. Source: Own elaboration.

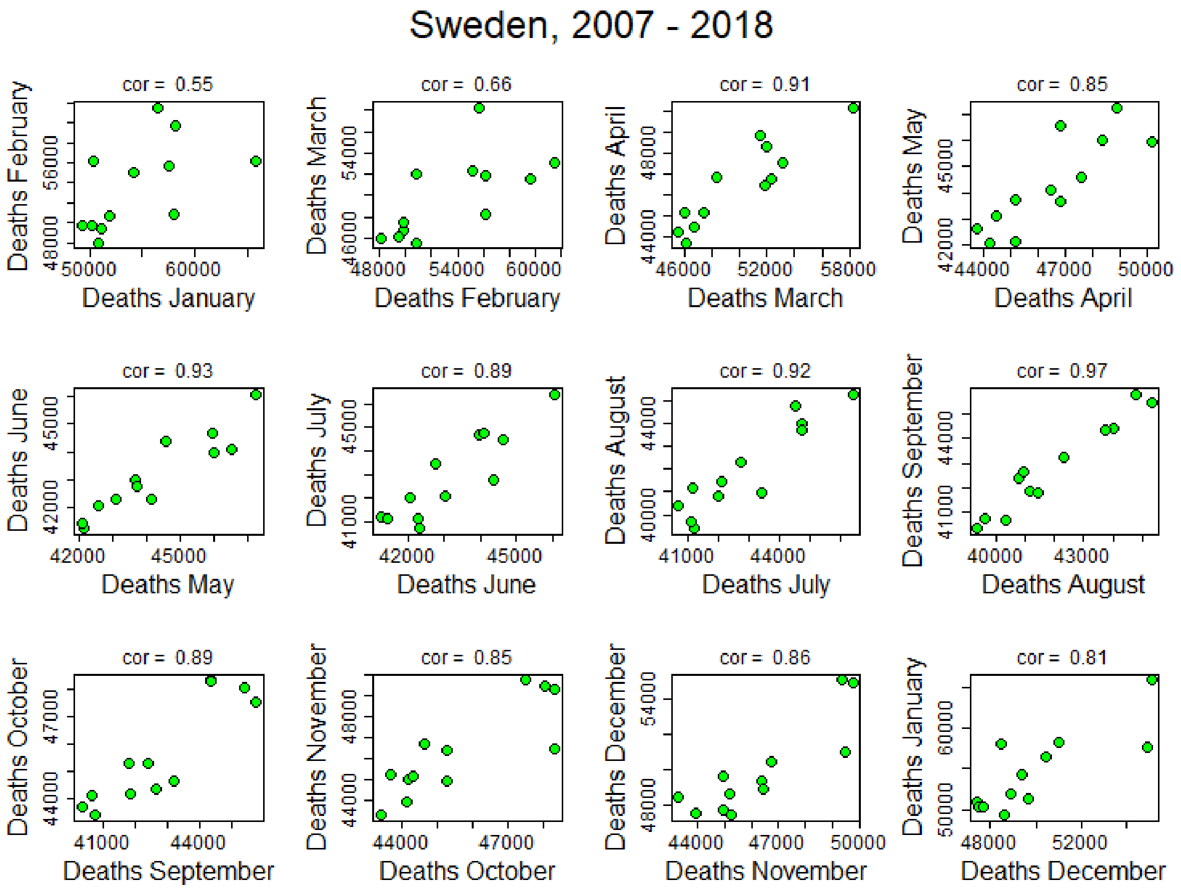


Supplementary Figure 4. Correlation between deaths in consecutive months, year 2007 to 2021, Sweden. Source: Own elaboration.

### 3. Supplementary Methods

#### Alternative forecasting methods

We compared our forecasts with two alternative forecasting methods: the 5-years-average and the Serfling model.

The Serfling model uses a periodic equation to model the monthly death counts ($D_t$)

$$D_t = \beta_1 + \beta_2 t + \gamma_1 \cos\frac{2\pi t}{n} + \delta_1 \sin\frac{2\pi t}{n} + \gamma_2 \cos\frac{4\pi t}{n} + \delta_2 \sin\frac{4\pi t}{n} + \varepsilon_t$$

with $n$ = *12* when monthly data were analyzed. In this model, the mortality rate depends on time *t* through a constant term $\beta_1$, a secular trend $\beta_2$, annual predictors $\gamma_1$and $\delta_1$, semiannual predictors $\gamma_2$ and $\delta_2$and an error term $\varepsilon_t$. The original paper by Serfling used 4-week periods, so the period denominator is $n$ = *13*. The unknown parameters are estimated at maximum likelihood from the fit of a linear regression to data from a 'training' period.

In our application, we assumed that the death counts are distributed following a quasi-Poisson, which allows to account for overdispersion.

### 4. Supplementary Results

#### Series of later/earlier ratios

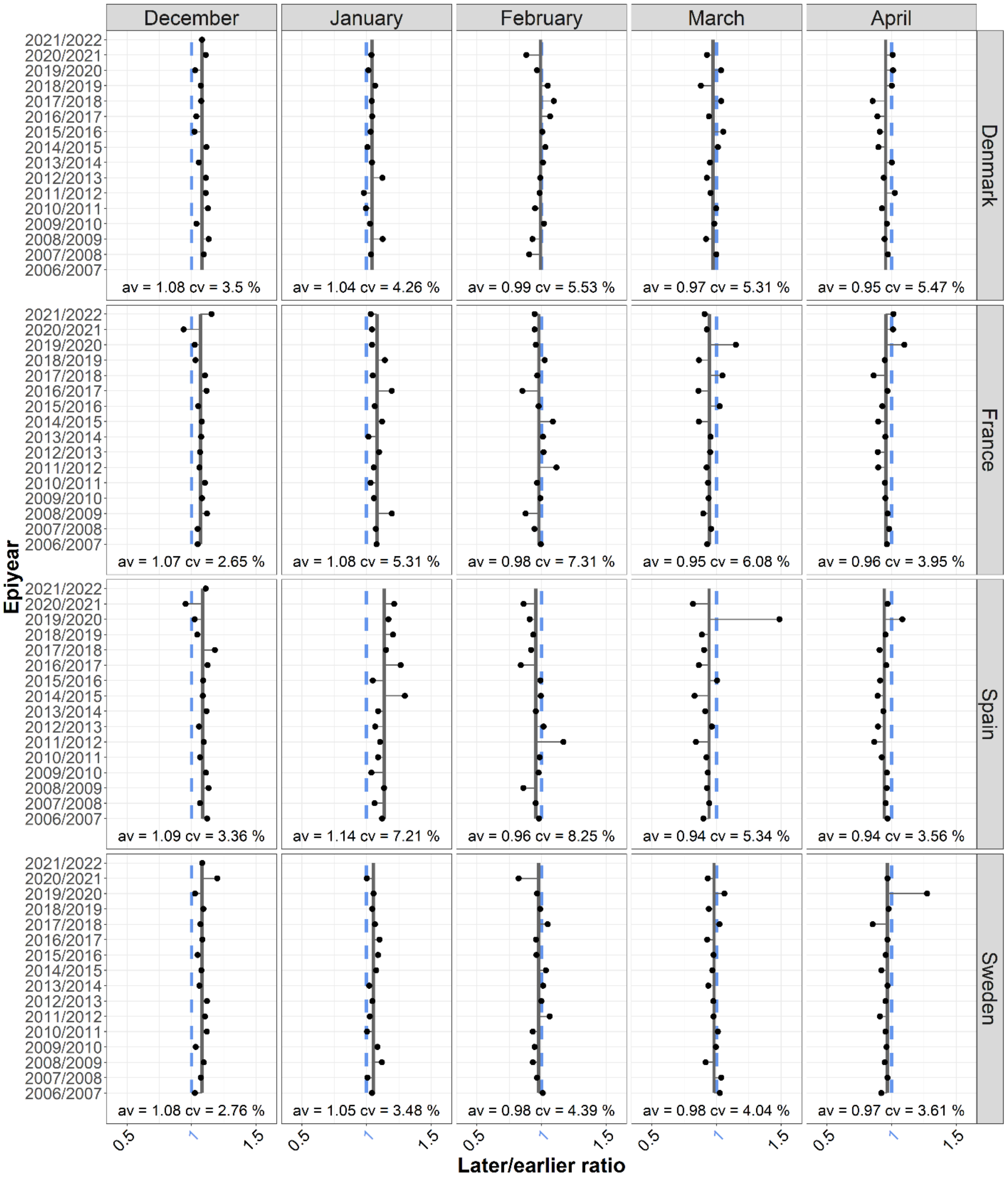


Supplementary Figure 5: Later/earlier ratios (black dots) and average later/earlier ratios (vertical black bars). Source: Own elaboration.

### 5. Supplementary Sensitivity Analysis

#### Sensitivity analysis of the series of later/earlier ratios

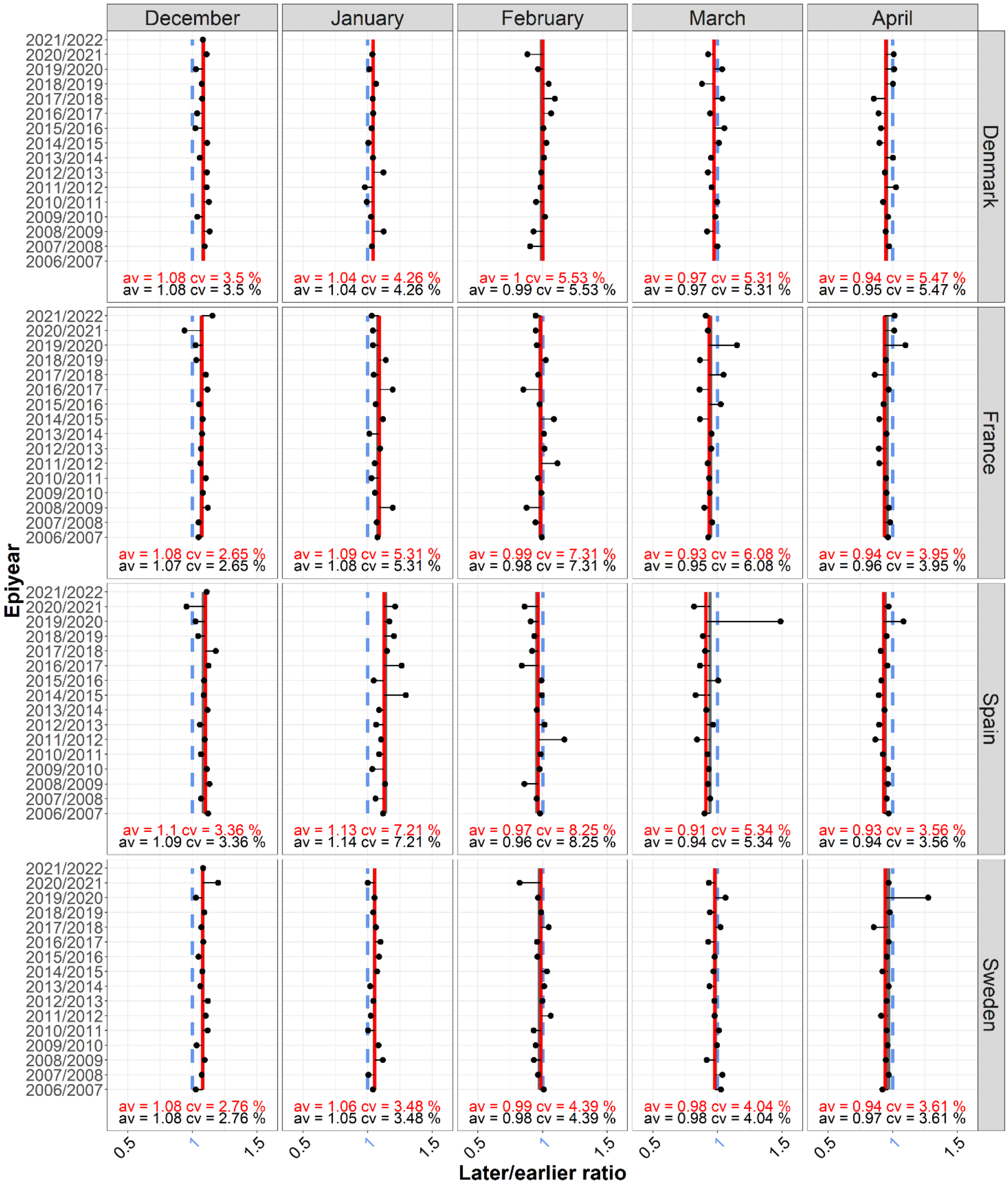

Supplementary Figure S6: Average later/earlier ratios with (vertical black bars) and without (vertical red bars) covid years, with a later period of one month and an earlier period of one month. Source: Own elaboration.
